# Accurate RR-interval extraction from single-lead, telehealth electrocardiogram signals

**DOI:** 10.1101/2025.03.10.25323655

**Authors:** Sharon Yuen Shan Ho, Zixuan Ding, David C. Wong, Florian Kristof, James Brimicombe, Martin R. Cowie, Andrew Dymond, Hannah Clair Lindén, Gregory Y. H. Lip, Kate Williams, Jonathan Mant, Peter H. Charlton

**Affiliations:** Department of Public Health and Primary Care, University of Cambridge, Cambridge, CB1 8RN, United Kingdom; TUM School of Computation, Information, and Technology, Technical University of Munich, Garching bei München, 85748, Germany; School of Cardiovascular Medicine & Sciences, Faculty of Lifesciences & Medicine, King’s College London, London, United Kingdom; Zenicor Medical Systems AB, Stockholm, 113 59, Sweden; Liverpool Centre for Cardiovascular Science at University of Liverpool, Liverpool John Moores University and Liverpool Heart & Chest Hospital, Liverpool, United Kingdom; Department of Clinical Medicine, Aalborg University, Aalborg, Denmark; Leeds Institute of Health Sciences, University of Leeds, Leeds, UK

**Author notes:** Email address:* (Sharon Yuen Shan Ho).

**Keywords:** atrial fibrillation, ECG, Electrocardiography, inter-beat interval, mhealth, mobile, signal processing, signal quality

## Abstract

Devices that record single-lead ECGs, such as smartwatches and handheld ECG recorders, hold promise for detecting undiagnosed atrial fibrillation (AF). Accurately extracting RR-intervals from telehealth ECGs is key for heart rhythm assessment. The aim of this study was to develop an algorithm to extract RR-intervals from telehealth ECGs, and assess whether the extracted RR-intervals are accurate and therefore suitable for analysis. Two datasets of 30-second handheld ECGs were used: TELE ECG Database (250 ECGs) and SAFER ECG dataset (507 ECGs). One of three high-performance primary QRS detectors, selected based on previous evidence, was used to detect QRS complexes and extract RR-intervals. These detections were compared to those from a secondary QRS detector to assess accuracy. All pairs of 3 primary and 18 secondary QRS detectors were tested. Accuracy was quantified using mean absolute error (MAE) and the proportion of time RR-intervals were assessed as accurate (coverage). Best performance was achieved using *unsw* and *nk* as primary and secondary detectors, with MAEs of 19.8ms and 16.3ms, and coverages of 89% on TELE and SAFER respectively. Using a single detector alone produced higher MAEs (23.8ms and 43.9ms on TELE; 38.2ms and 41.7ms on SAFER). Accuracy was similar between AF and non-AF, but reduced on low-quality signals (50.8 vs. 7.7ms, p*<*0.001). In conclusion, the recommended algorithm produced more accurate RR-intervals than using a single QRS detector, maintaining accuracy during AF, although accuracy was reduced on low-quality signals.

**Highlights:**

- Algorithm extracts RR-intervals from ECGs and assesses their accuracy
- Algorithm was developed using two datasets of ECGs collected using different devices
- The impacts of arrhythmia and noise on algorithm performance were assessed
- The algorithm uses a pair of openly available QRS detection algorithms

## 1. Introduction

Single-lead electrocardiogram (ECG) signals can now be recorded using mobile devices such as smartwatches [1] and handheld ECG recorders [2]. A promising application for such devices is to detect undiagnosed atrial fibrillation (AF), a common heart arrhythmia which confers an increased risk of stroke [3], and yet is often undiagnosed [4]. Indeed, it is feasible to use hand-held ECG recorders in population-level AF screening of older adults [5, 6]. The large-scale use of such devices generates too many ECGs for them all to be manually reviewed, particularly when acquiring multiple ECGs per participant to detect paroxysmal AF. Consequently, automated algorithms are now commonly used to identify ECGs which show possible signs of AF and warrant clinical review [7].

RR-intervals are widely used in ECG analysis, with routine applications including heart rate estimation [8], heart rate variability analyses [9], and arrhythmia detection [10]. Our current interest is in using RR-intervals to assess the level of irregularity of a heart rhythm and to identify ECGs showing signs of AF [11]. RR-intervals can be automatically extracted from ECGs by using a QRS detection algorithm [12], followed by calculating the intervals between consecutive R-waves [13]. Statistical methods or machine learning models can then be used to determine whether a set of RR-intervals shows an irregular pattern consistent with AF [10]. Consequently, it is important to develop algorithms to extract RR-intervals accurately from ECG signals. This task is more challenging with telehealth ECGs than ECGs collected under clinical supervision, as telehealth ECG signals are more prone to noise [14], and often contain only a single lead, preventing the use of multiple leads for accurate QRS detection [15]. Consequently, QRS detection algorithms may be less accurate when used with telehealth ECGs [12], leading to errors in extracted RR-intervals.

In this study we present a novel algorithm to extract RR-intervals from single-lead, telehealth ECGs, and automatically assess the accuracy of the extracted RR-intervals. The algorithm extracts RR-intervals using a primary QRS detector, and automatically assesses their accuracy by comparing the QRS detections with those provided by a secondary QRS detector. RR-intervals for which both QRS detectors agreed on QRS detections are assessed as accurate, and RR-intervals containing any disagreement on QRS detections are assessed as inaccurate. The main novelty is the development of an algorithm which automatically assesses the accuracy of extracted RR-intervals. Whilst many ECG signal quality assessment algorithms have been developed in the past [16, 17, 18, 19, 20, 21], we are not aware of an algorithm specifically designed to assess the accuracy of extracted RR-intervals from the underlying ECG signal.

We hypothesised that the novel algorithm would provide more accurate RR-intervals than using a single QRS detector alone. We addressed the following research questions:

1. Does using a pair of QRS detectors improve performance beyond using a single QRS detector?
2. Which combination of QRS detectors provides the most accurate RR-intervals in two use cases: (i) when seeking the highest accuracy regardless of the proportion of RR-intervals assessed to be accurate; and (ii) when seeking the highest accuracy when at least a threshold proportion of RR-intervals are assessed to be accurate?
3. How do atrial fibrillation and signal quality influence algorithm performance?

The main contributions to knowledge were: (i) identifying the most suitable QRS detectors for use in the algorithm; (ii) establishing the performance of the algorithm on real-world telehealth ECGs; and (iii) providing an open-source Python implementation of the algorithm. To achieve these, we performed a systematic evaluation of different combinations of 18 QRS detectors on two datasets of real-world telehealth ECGs. This work extends work presented at a conference [22] by using an additional dataset of ECGs collected during screening for AF, containing arrhythmias and noise. Some material has been reproduced or adapted from [22] under the CC BY 4.0 licence.

## 2. Methods

### 2.1. Datasets

In this study we used two datasets of telehealth single-lead ECGs: the publicly available TELE ECG Database (TELE), and ECGs from the SAFER AF Screening Programme (SAFER). In both cases ECGs were recorded at home, from the hands (specifically, using the thumbs in the case of SAFER), using devices with dry electrodes. Both datasets contain 30-second ECG signals sampled at 500Hz, as well as manual annotations of QRS complexes. These datasets are now described.

#### 2.1.1. TELE ECG Database (TELE)

As described in [14, 23], the TELE ECG Database contains 250 ECGs recorded by home-dwelling patients suffering from chronic obstructive pulmonary disease and/or congestive heart failure. ECGs were recorded between the hands using the TeleMedCare Health Monitor (TeleMedCare Pty Ltd Sydney, Australia). The dataset includes 29 ECGs which were specifically selected as they were deemed to be poor quality [14]. One ECG lasted longer than 30s and was truncated to 30s for this study [12].

#### 2.1.2. SAFER ECG Dataset (SAFER)

The SAFER ECG dataset contains 507 ECGs. A subset of this dataset, containing 479 ECGs was reported in [12]. For the present study a further 28 low-quality ECGs were added. The ECGs were recorded by older adults aged 65 and over during the SAFER (Screening for Atrial Fibrillation with ECG to Reduce stroke) research programme (ISRCTN 16939438). The ECGs used in this study were collected in the SAFER Feasibility Study, which has been described in [6]. Briefly, ECGs were recorded between the thumbs using the Zenicor ONE device (Zenicor Medical Systems AB, Sweden). The dataset includes: 183 high-quality and 28 low-quality ECGs exhibiting AF (denoted SAFER-AF-HIGH and SAFER-AF-LOW respectively); and 199 high-quality and 97 low-quality ECGs from subjects without AF (SAFER nonAF-HIGH and SAFER-nonAF-LOW respectively). Quality labels were generated using the Cardiolund ECG Parser algorithm (Cardiolund AB) [7], and the presence of AF was determined by clinicians with the aid of the Cardiolund algorithm as described in [24, 12].

The SAFER Feasibility Study in which the SAFER ECG dataset was acquired was approved by the London Central NHS Research Ethics Committee (18/LO/2066). All participants gave written informed consent to participate in the study. The study was conducted in accordance with the Declaration of Helsinki.

### 2.2. QRS Detection

We used three high-performance QRS detection algorithms to detect QRS complexes and extract RR-intervals. These were the Neurokit (*nk*) [25, 26], two-average (*two-avg*) [27], and University of New South Wales (*unsw*) [14] algorithms. These have previously been found to perform well on the TELE and SAFER datasets in [12], achieving *F*_1_ scores of *≥*0.96 on high-quality ECGs, and *≥*0.70 on low-quality ECGs. The original *unsw* implementation was written in MATLAB. For this study we translated *unsw* into Python, and report results obtained using the new Python version in the main text (with corresponding results for the original MATLAB implementation in an appendix). To account for primary QRS detectors not providing the precise locations of R-wave peaks, each detected R-wave location was refined by identifying the point of highest amplitude within *±* 50ms of the initial R-wave detection.

A total of 18 QRS detection algorithms were used to assess the accuracy of extracted RR-intervals. These consisted of: (i) the three aforementioned algorithms; (ii) 13 of the 15 remaining algorithms used in a recent study of open-source QRS detectors [12] (two algorithms denoted *gamb* and *mart* were excluded due to their poor performance in [12]); and (iii) two further algorithms included to ensure all of the top-performing QRS detectors in a comparison study [28] were included: the U3 transform algorithm (*u3*) [29] and the difference operation algorithm (*dom*) [30]. All algorithms were implemented in Python, except *jqrs*, *rdeco*, and *rpeak*, which were implemented in MATLAB.

### 2.3. RR-Interval Extraction and Accuracy Assessment

The process for RR-interval extraction and accuracy assessment is summarised in Figure 1 and now described.

**Figure 1:**
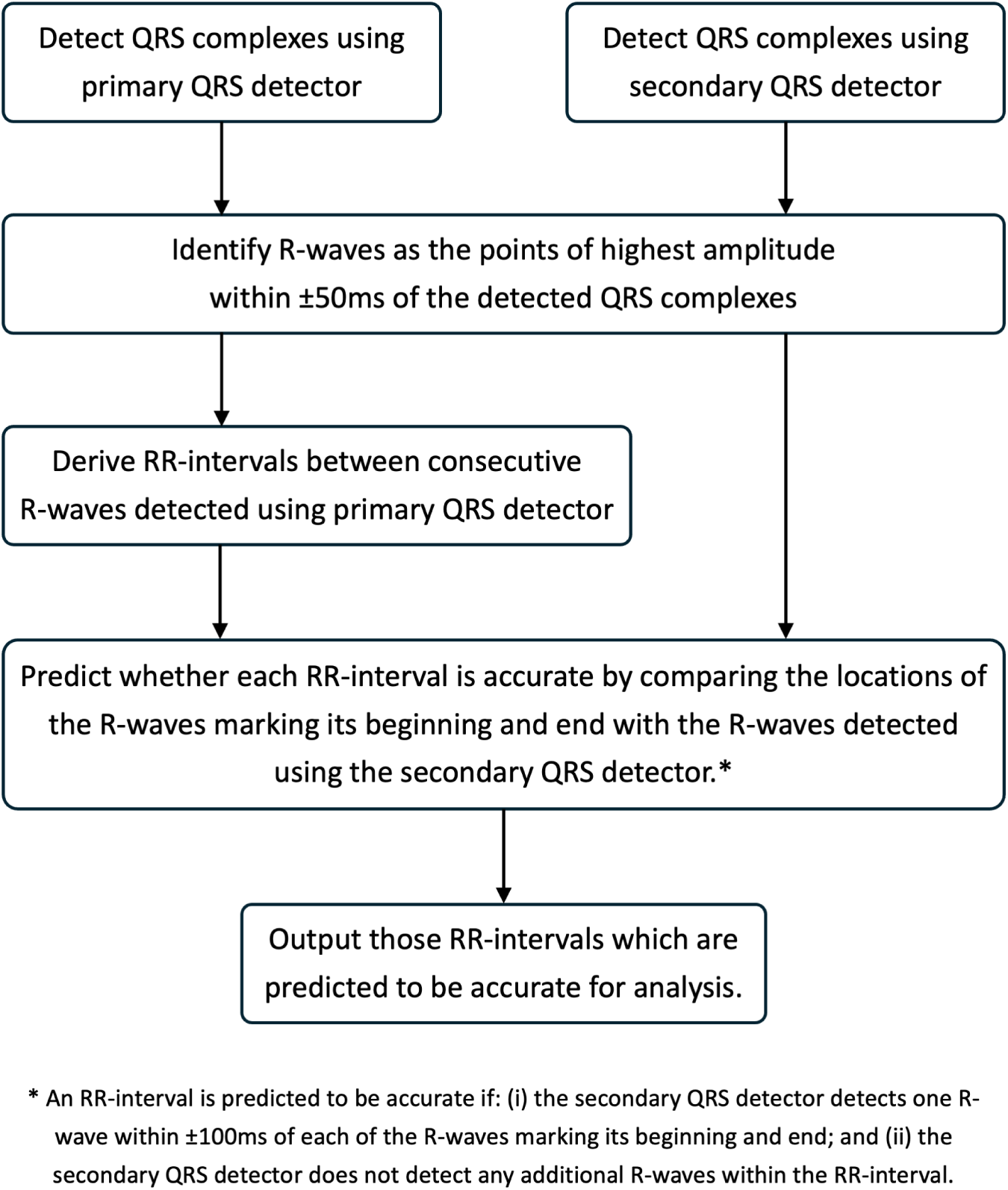
Algorithm for extracting RR-intervals and assessing whether they are accurate.

RR-intervals were extracted using a single, high-performance ‘primary’ QRS detector (*i.e. nk*, *two-avg*, or *unsw*). To do so, QRS complexes were detected, and then RR-intervals were calculated as the time delays between consecutive QRS complexes. Next, the accuracy of each RR-interval was assessed by comparing the locations of the QRS complexes marking its beginning and end with those detected by a secondary detector (which could be any one of the 18 QRS detectors). Briefly, if the two detectors agreed on the locations of both the QRS complexes, then that RR-interval was assessed to be accurate, as demonstrated in Figure 2. To give further detail: an RR-interval was deemed to be accurate if the QRS complexes marking its beginning and end were detected by both the primary and secondary detector, and no further QRS complexes were detected by either detector within the RR-interval. When assessing whether a QRS complex was detected by both the primary and secondary detector, we allowed a tolerance of *±* 100ms, *i.e.* if the secondary detector detected a QRS complex within 100ms either side of the time of that detected by the primary detector, then this QRS complex was deemed to have been detected by both detectors. A value of *±* 100ms was chosen as a compromise between lower values which result in higher accuracy, and higher values which result in RR-intervals being assessed to be accurate for a higher proportion of the time. The preliminary analysis which informed this decision is shown in Appendix A.

**Figure 2:**
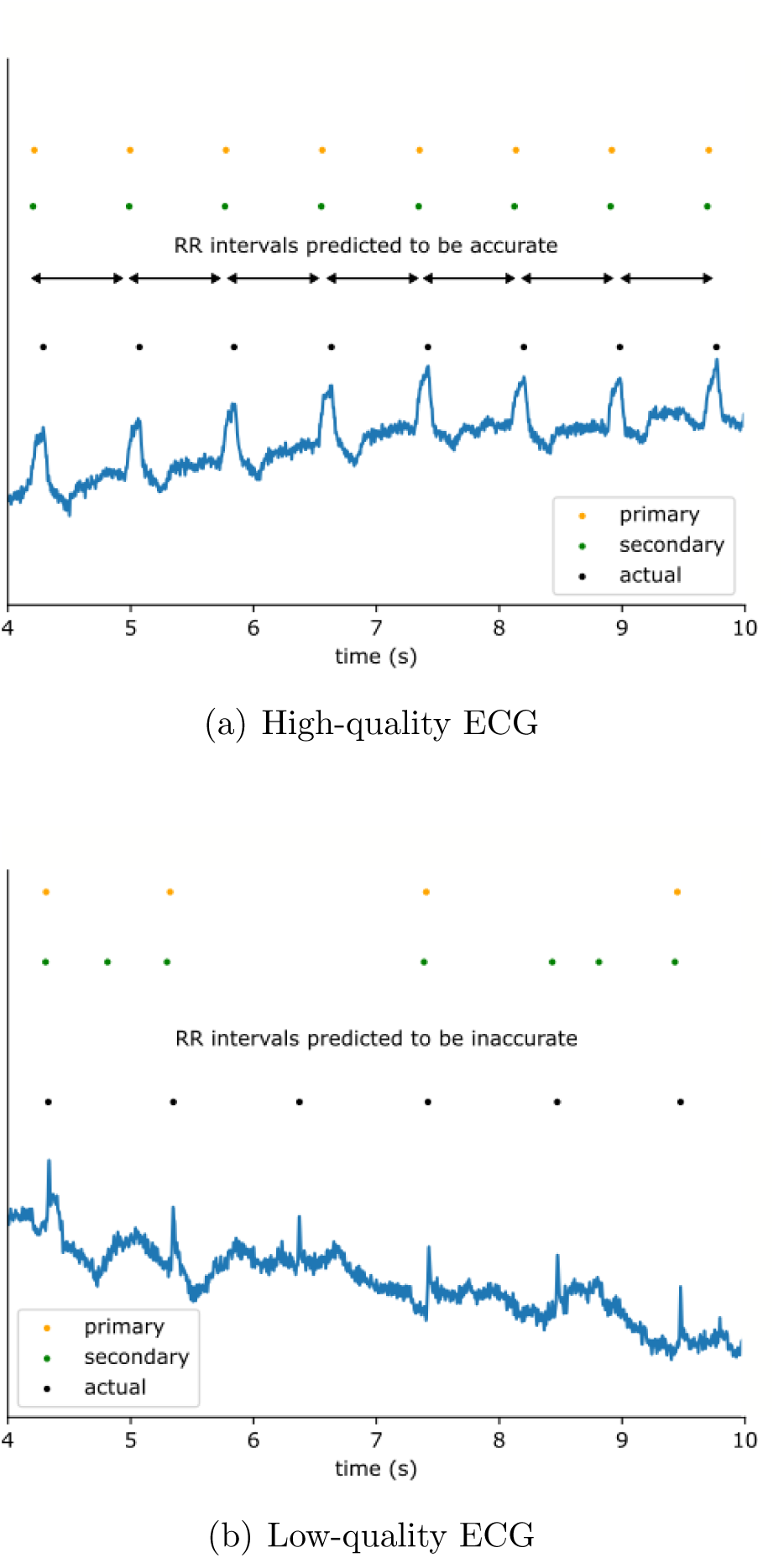
Using two QRS detectors to assess the accuracy of RR-intervals. RR-intervals were assessed to be accurate if the primary and secondary detections were in agreement. (a) shows an example where the detectors were in agreement, (b) shows an example where they disagreed. For comparison, the actual manual annotations of QRS complexes are shown in black.

The proposed approach for assessing the accuracy of RR-intervals is a modification of the *bSQI* approach to ECG signal quality assessment [31]. The *bSQI* approach provides a quantitative assessment of signal quality, defined as “a sliding-window ratio of consensus beats to total beats (consensus and non-consensus)” [31]. This approach has been used in several studies [32, 16, 33, 21, 34, 35, 36, 37, 28], albeit possibly with slight variations in the calculation (such as calculating the ratio of consensus beats to total beats detected by one beat detector [16, 37]). In this work we modified the original approach to produce a binary classification for each RR-interval, rather than a quantitative assessment for a window of ECG signal (*e.g.* a 10-second window).

RR-intervals were extracted and their accuracy assessed for each combination of the three primary detectors and 18 secondary detectors, resulting in 54 pairs of QRS detectors. However, we note that the three combinations which used the same primary and secondary detectors are equivalent to using a single QRS detector with no accuracy assessment.

### 2.4. Performance Evaluation

The performance of the algorithm when using each pair of QRS detectors was assessed as follows. A set of RR-intervals, alongside algorithm assessments of whether each RR-interval was accurate or not, were obtained for each ECG and each pair of QRS detectors. In addition, reference RR-intervals were calculated as the time delays between manually annotated QRS complexes.

To facilitate comparison, the reference and algorithm-extracted time series of RR-intervals were interpolated (using sample and hold interpolation) at 500 Hz at the same points in time. Any time points for which either time series was not available were discarded, thus ensuring the two time series had the same start and end times. To account for un-annotated portions of ECG signals, any periods with reference RR-intervals of *>*3 seconds were excluded from the analysis.

Performance was assessed using two metrics: (i) the proportion of time for which RR-intervals were assessed to be accurate, and therefore would be available for analysis (known as the ‘coverage’); and (ii) the mean absolute error (MAE) between reference and extracted RR-intervals (in ms). Each metric was calculated for each ECG. Then the ‘mean coverage’ and the ‘mean MAE’ were calculated for each pair of QRS detectors on each dataset. The calculation for MAE was based solely on periods of time in which the algorithm assessed that the extracted RR-intervals were accurate, mimicking the intended real-world use of the algorithm, where only those RR-intervals assessed to be accurate would be analysed.

### 2.5. Identifying the Best-Performing Pairs of QRS Detectors

The best-performing pairs of QRS detectors were identified for two use cases. In the first use case, we identified the pair with the lowest mean MAE. This approach prioritises the accuracy of RR-intervals, at the expense of potentially identifying pairs of QRS detectors which may make a lower proportion of RR-intervals available for analysis. The pair identified using this approach could be suitable in scenarios where ECG data are plentiful and it is acceptable to discard a high proportion of RR-intervals. In the second use case, we identified the pair with the lowest mean MAE whilst achieving a sufficiently high mean coverage (chosen to be *≥*85%). The pair identified using this approach could be suitable in scenarios where accuracy requirements are not as strict, and greater priority is placed on retaining a higher proportion of RR-intervals.

The following subgroup analyses were performed. First, we investigated how performance varied between high and low quality signals by comparing performance between the SAFER nonAF-HIGH (high-quality) and SAFER nonAF-LOW (low-quality) subgroups. Second, we investigated how performance varied in the presence of arrhythmia by considering the SAFER AF-HIGH (AF present) and SAFER nonAF-HIGH (AF not present) subgroups. The MAE and coverage when using each pair of QRS detectors were compared between subgroups using the Mann-Whitney U test with a significance level of *p* = 0.05.

## 3. Results

### 3.1. Overall performance

The results for the mean MAE and mean coverage (proportion of time for which the algorithm assessed RR-intervals would be accurate) on the TELE and SAFER datasets are presented in Figure 3.

Figure 3(a) and 3(c) show that there was a wide range of mean MAEs when using different pairs of QRS detectors, ranging from 16ms to 86ms on the TELE dataset, and 12ms to 193ms on the SAFER dataset. Furthermore, it shows that RR-intervals were generally more accurate (lower mean MAEs) when using *unsw* or *nk* as the primary QRS detector, compared to using *two-avg*. There were minimal differences in accuracy when using *unsw* or *nk* as the primary QRS detector: the only significant differences were when the primary and secondary detectors were the same (*e.g.* [*nk*, *nk*]), or when using *dom* on SAFER.

**Figure 3:**
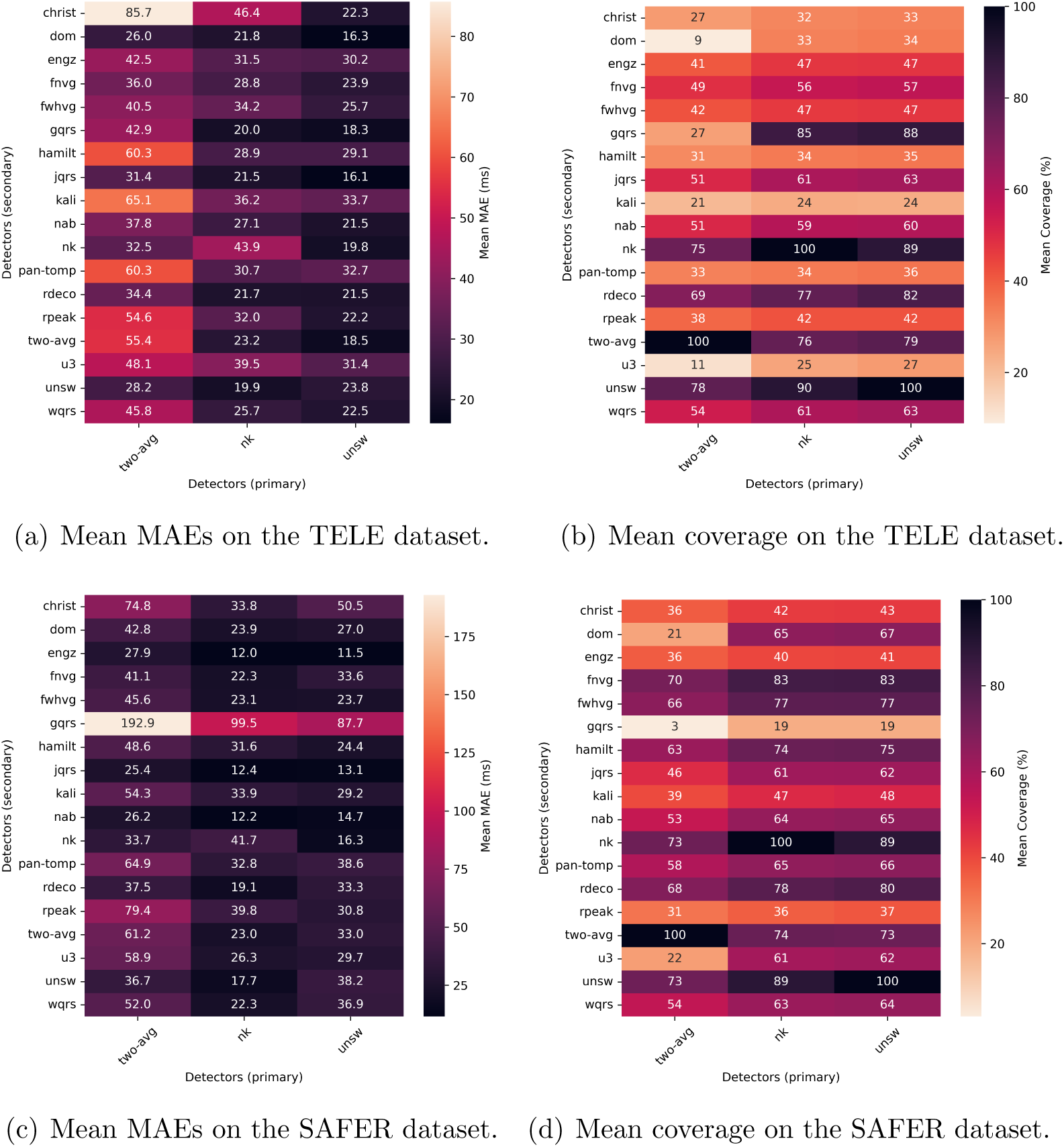
Overall performance. Heatmaps showing the mean MAE ((a) and (c)) and mean coverage ((b) and (d)) for each pair of QRS detectors on the TELE ((a) and (b)) and SAFER ((c) and (d)) datasets. Here, the ‘coverage’ is the proportion of time for which RR-intervals were assessed to be accurate.

MAEs were generally lower when using a pair of QRS detectors to exclude RR-intervals assessed to be inaccurate, in comparison to using a single QRS detector without accuracy assessment. For instance, the [*nk*, *unsw*] pair produced significantly lower mean MAEs of 19.9 and 17.7ms on TELE and SAFER datasets, compared to MAEs of 43.9 and 41.7ms when using [*nk*, *nk*] (*i.e.* using *nk* alone without accuracy assessment) (both p*<*0.001). Similarly, mean MAEs were significantly lower for [*unsw*, *nk*] than [*unsw*, *unsw*] on SAFER (p*<*0.001), although there was no significant difference on TELE.

Figures 3(b) and 3(d) show that there was a wide range of proportions of time for which RR-intervals were assessed to be accurate, ranging from 9 to 90% on the TELE dataset, and 3 to 89% on the SAFER dataset (ignoring pairs of the same QRS detectors).

The results shown in this section were obtained using the new Python implementation of *unsw*. The corresponding results for the original MATLAB implementation were similar, and are provided in Appendix B.

### 3.2. Best-performing pairs of QRS detectors

Figure 4 illustrates the relative performance of pairs of QRS detectors when considering both their mean MAE and mean coverage. Each point represents a pair of different QRS detectors (where pairs of the same QRS detectors have been removed).

**Figure 4:**
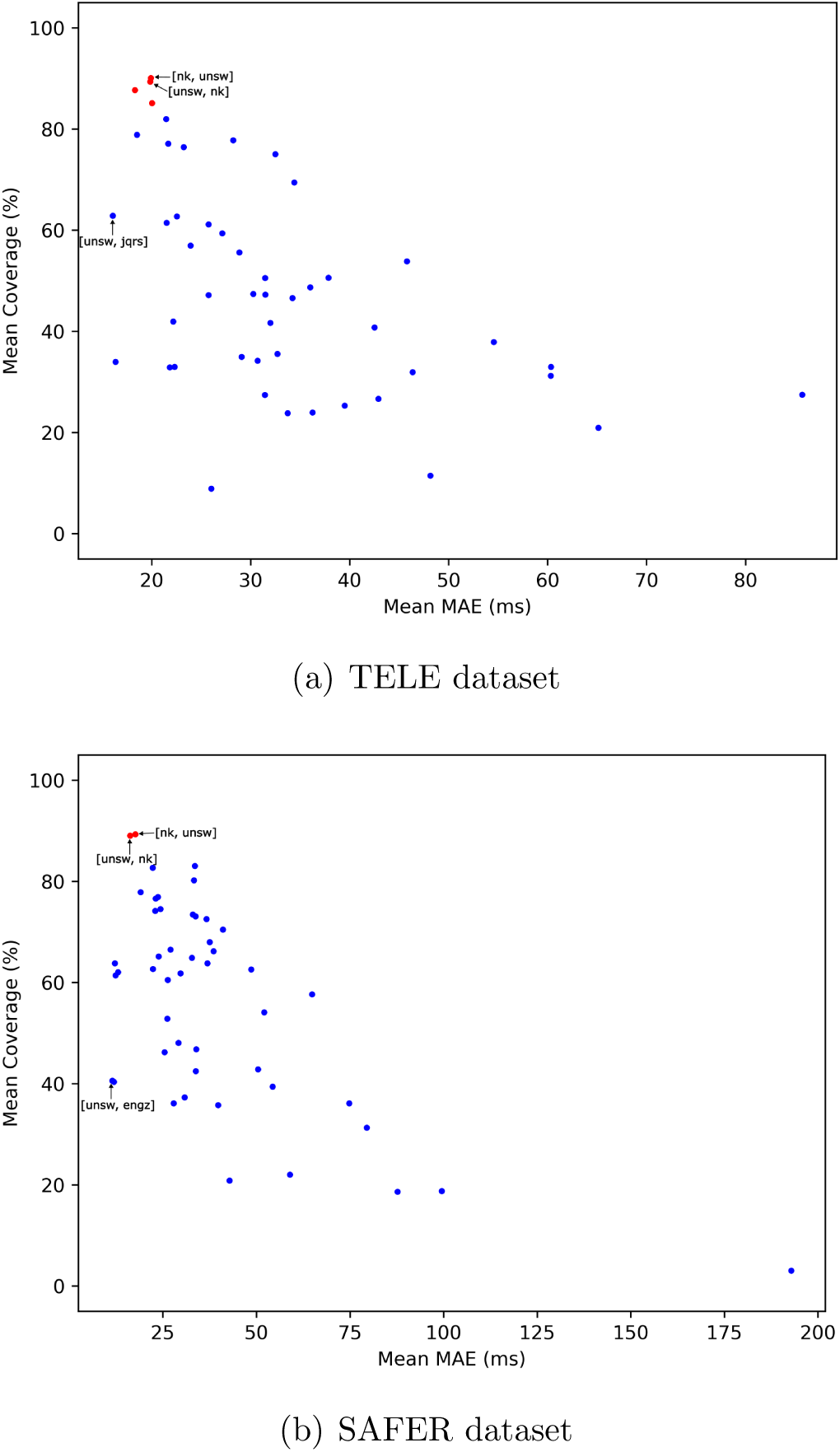
Identifying the best-performing pairs. Scatter plots showing the mean coverage against the mean MAE for all pairs of different QRS detectors on: (a) the TELE dataset, and (b) the SAFER dataset. Those pairs of QRS detectors which achieved *≥*85% coverage are marked in red. The recommended pairs and the pair with the lowest mean MAE are annotated.

#### 3.2.1. Pairs of QRS detectors producing the lowest MAEs

The most accurate RR-intervals were obtained on the TELE dataset when using [*unsw*, *jqrs*] as [primary, secondary] QRS detectors, with a mean MAE of 16.1ms; and on the SAFER dataset when using [*unsw*, *engz*], with a mean MAE of 11.5ms. Whilst these pairs provided the lowest mean MAEs, they assessed that only 63% and 41% of RR-intervals were accurate respectively. In contrast, many other pairs of QRS detectors achieved a much higher mean coverage, with only a slight increase in mean MAE (see Figure 4).

#### 3.2.2. Pairs of QRS detectors producing the lowest MAEs and sufficient coverage

Considering our second approach to selecting the best-performing pair of QRS detectors: the most suitable pairs are those in the top left corners of the scatter plots in Figure 4, *i.e.* those pairs which assessed a high proportion of RR-intervals to be accurate, and achieved a low mean MAE. Our requirement of providing *≥* 85% coverage was achieved on the TELE dataset when using: *nk* or *unsw* as the primary QRS detector, and *gqrs*, *nk* or *unsw* as the secondary QRS detector. Of these, [*unsw*, *gqrs*] was the best-performing pair, followed by [*unsw*, *nk*]. These assessed RR-intervals to be accurate for 88% and 89% of the time, and achieved mean MAEs of 18.3ms and 19.8ms respectively. On the SAFER dataset, the requirement of *≥* 85% coverage was only achieved when using either combination of *nk* and *unsw*. The best performing pair was [*unsw*, *nk*], followed by [*nk*, *unsw*], both achieving a coverage of 89%, and achieving mean MAEs of 16.3ms and 17.7ms respectively.

### 3.3. Factors influencing algorithm performance

The impact of factors on algorithm performance was investigated by comparing performance across the SAFER subgroups: AF-HIGH (high quality ECGs exhibiting AF); nonAF-HIGH (high quality ECGs not exhibiting AF); and nonAF-LOW (low quality ECGs not exhibiting AF). Figure 5 provides numerical results for the mean MAE and mean coverage of each pair of QRS detectors on each subgroup. Figure 6 shows scatter plots of mean coverage against mean MAE for each subgroup.

**Figure 5:**
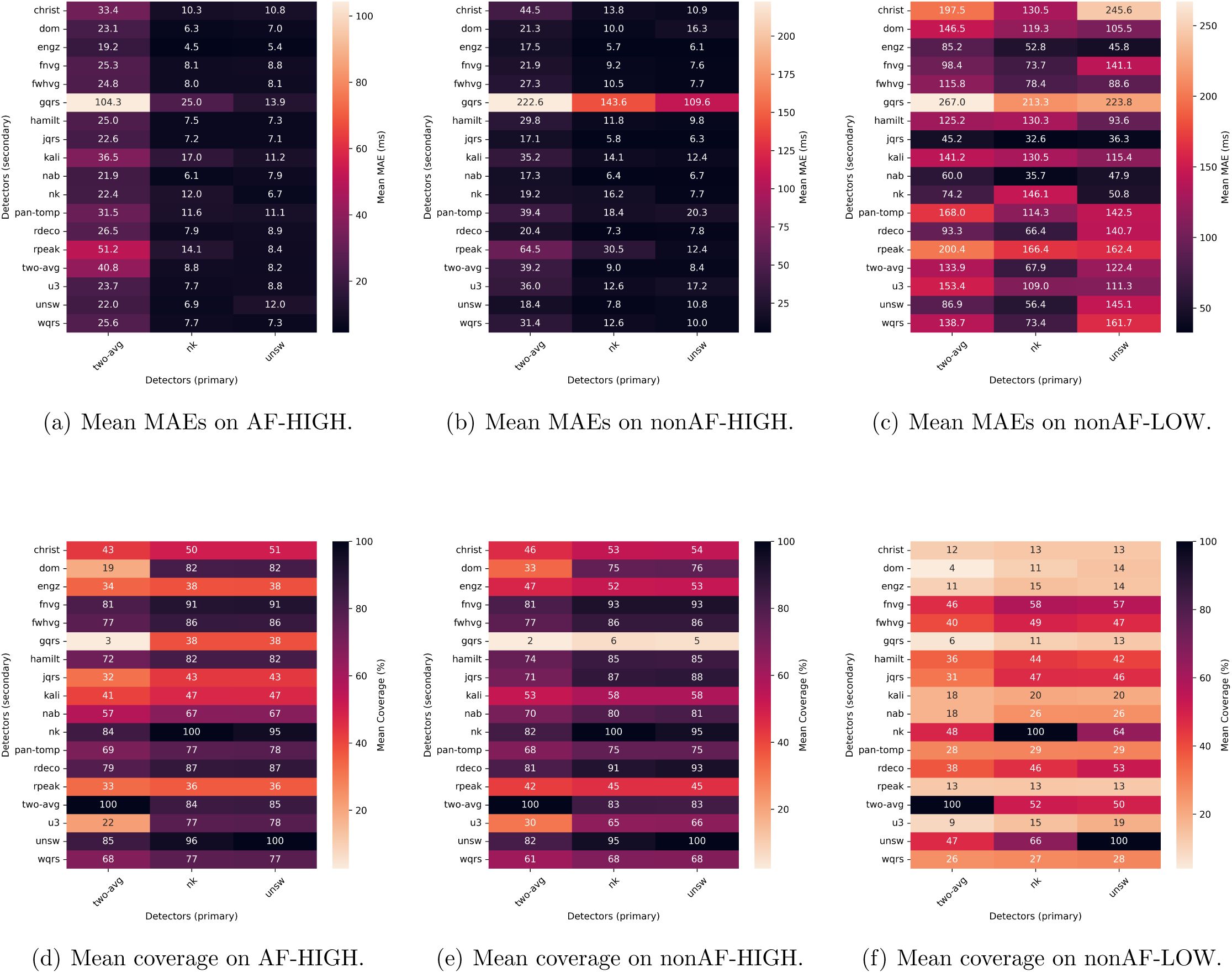
Comparing performance across SAFER subgroups. Heatmaps showing the mean MAE ((a), (b) and (c) and mean coverage ((d), (e) and (f)) for each pair of QRS detectors on the SAFER AF-HIGH ((a) and (d)), SAFER nonAF-HIGH ((b) and (e)), and SAFER nonAF-LOW ((c) and (f)) subgroups.

**Figure 6:**
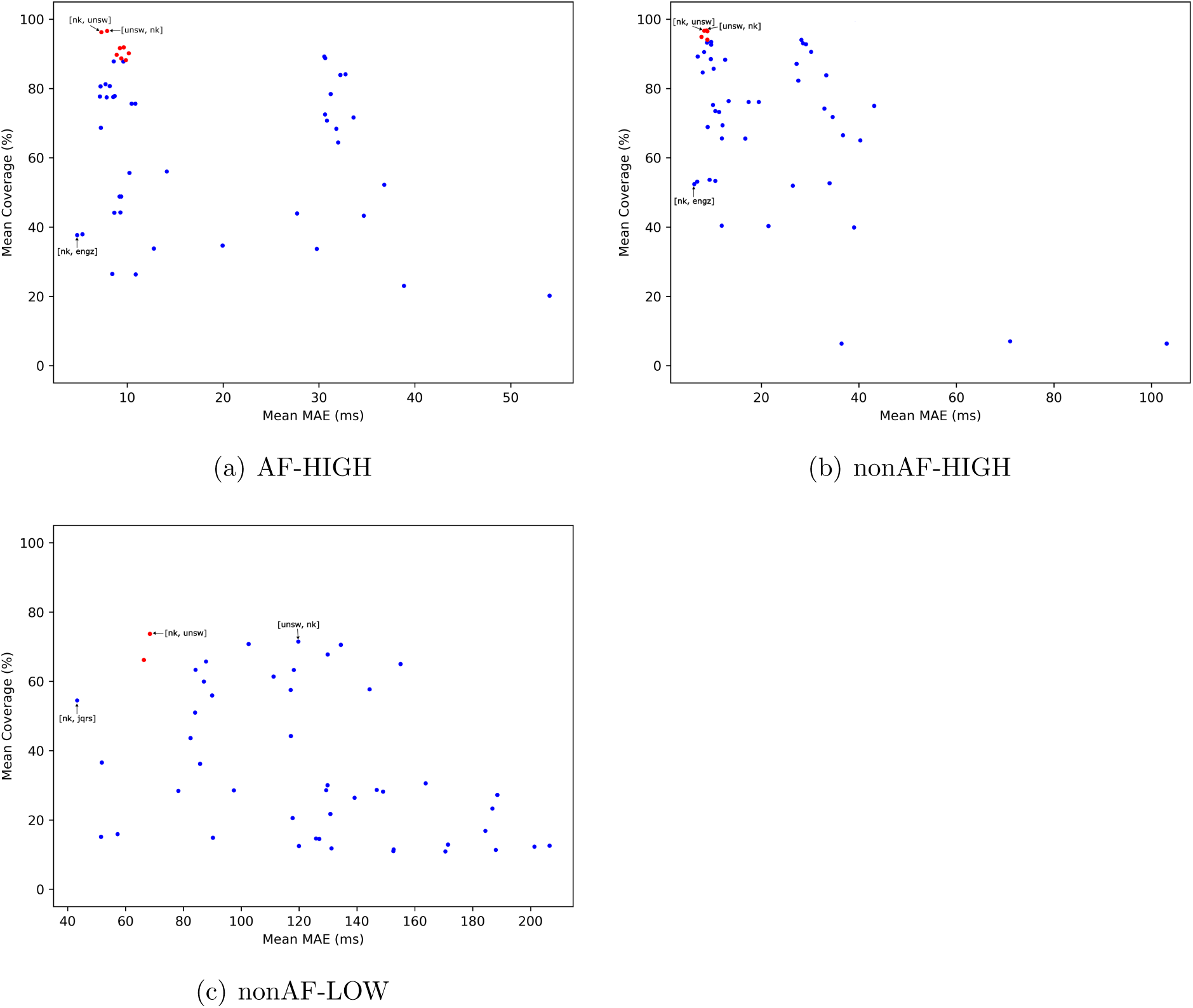
Identifying the best-performing pairs on SAFER subgroups. Scatter plots showing the mean coverage against the mean MAE for all pairs of different QRS detectors on (a) SAFER AF-HIGH, (b) SAFER-nonAF-HIGH, and (c) SAFER-nonAF-LOW. Those pairs of QRS detectors which achieved *≥*85% coverage (or *≥*65% on SAFER-nonAF-LOW) and lower MAEs are marked in red. The recommended pairs and the pair with the lowest mean MAE are annotated.

#### 3.3.1. The impact of atrial fibrillation on algorithm performance

Considering the impact of atrial fibrillation, mean MAEs on SAFER AF-HIGH and SAFER nonAF-HIGH are shown in Figure 5(a) and 5(b) respectively. The previously identified best-performing pairs which produced a mean coverage of *≥*89% on the entire dataset, [*nk*, *unsw*] and [*unsw*, *nk*], achieved MAEs of 6.9ms and 6.7ms on AF-HIGH respectively, and 7.8ms and 7.7ms on nonAF-HIGH respectively, and achieved mean coverages of 96 and 95% on AF-HIGH and 95% for both on nonAF-HIGH. There was a significant difference between MAEs on AF-HIGH and nonAF-HIGH sub-groups for 26 out of 54 pairs, of which the mean MAEs were higher in AF for 16 of the 26 pairs. However, there was no significant difference in MAE between AF and non-AF for either of the recommended [*unsw*, *nk*] or [*nk*, *unsw*] pairs. Similarly, there was a significant difference between coverages on AF-HIGH and nonAF-HIGH subgroups for 24 out of 54 pairs, of which the mean coverages were higher in non-AF for 17 out the 24 pairs. Similarly, there was no significant difference in coverage between AF and non-AF for either of the recommended [*unsw*, *nk*] and [*nk*, *unsw*] pairs.

Figures 6(a) and 6(b) illustrate that the best-performing pairs on SAFER AF-HIGH and SAFER nonAF-HIGH remained the same as those identified on the entire SAFER dataset: [*nk*, *unsw*] followed by [*unsw*, *nk*].

#### 3.3.2. The impact of signal quality on algorithm performance

Considering the impact of signal quality, mean MAEs on SAFER nonAF-HIGH and SAFER nonAF-LOW are shown in Figure 5(b) and 5(c) respectively. MAEs were much higher on low quality ECGs (nonAF-LOW) than high quality ECGs (nonAF-HIGH). For instance, the lowest mean MAE was 5.7ms on SAFER nonAF-HIGH, compared to 32.6ms on SAFER nonAF-LOW. Mean MAEs were significantly higher in nonAF-LOW compared to nonAF-HIGH for all 54 pairs.

The proportion of time for which RR-intervals were assessed to be accurate (the mean coverage) was much lower on low quality ECGs. The highest proportions were achieved by the previously identified best-performing pairs, [*nk*, *unsw*] and [*unsw*, *nk*], which achieved mean coverages of 66% and 64% resepectively on nonAF-LOW compared to 95% for both on nonAF-HIGH. There was a significant difference between coverages on nonAF-HIGH and nonAF-LOW for 51 out of 54 pairs, of which the mean coverages were higher on nonAF-HIGH than nonAF-LOW for 48 out of the 51 pairs.

Figure 6(c) illustrates that the best-performing pairs on SAFER nonAF-LOW differed slightly from those identified on the entire SAFER dataset: [*nk*, *two-avg*] followed by [*nk*, *unsw*], with [*unsw*, *nk*] having a higher mean MAE.

## 4. Discussion

### 4.1. Summary of Findings

This study demonstrates the feasibility of extracting RR-intervals accurately from telehealth ECG signals using a pair of QRS detectors. The main findings were as follows. First, as hypothesised, the accuracy of RR-intervals increased when using a pair of QRS detectors to exclude RR-intervals deemed to be inaccurate, in comparison to using a single QRS detector and assuming that all RR-intervals are accurate. Second, the [*unsw*, *nk*] combination of QRS detectors was found to provide the best performance. Third, the approach performed well during AF, with no significant differences in performance between AF and non-AF for the recommended QRS detector pairs. Fourth, the approach greatly improved the accuracy of RR-intervals derived from low-quality ECGs in comparison to using a single QRS detector alone. However, RR-intervals derived from low-quality ECGs were not as accurate as those extracted from high-quality ECGs.

This approach could be used to automatically exclude inaccurate RR-intervals from analysis. We observed a trade-off between the accuracy of RR-intervals and the proportion of time for which they were assessed to be accurate: the highest accuracy was obtained when using pairs of QRS detectors which frequently differed in their QRS detections, sometimes resulting in RR-intervals being available for less than half of the time. On the other hand, some pairs of QRS detectors resulted in RR-intervals being available for the vast majority of the time, whilst still achieving reasonable accuracy. For instance, the recommended [*unsw*, *nk*] combination of QRS detectors achieved MAEs of *<*20ms across two datasets compared to 23.8ms and 38.2ms when using the a single detector alone. This came at the expense of discarding RR-intervals for only 11% of the time.

### 4.2. Recommended Algorithms

When requiring accurate RR-intervals for a high proportion of the time, we recommend *unsw* as the primary QRS detector, and *nk* as the secondary QRS detector (denoted as [*unsw*, *nk*]). [*unsw*, *nk*] achieved a mean MAE of *<*20ms and mean coverage of 89% across both datasets and all SAFER subgroups except low quality SAFER ECGs. The order of the *nk* and *unsw* QRS detectors had little impact on performance, and the [*unsw*, *nk*] and [*nk*, *unsw*] combinations were among the top three performing combinations across both datasets. However, we note a higher MAE for [*unsw*, *nk*] than [*nk*, *unsw*] on SAFER nonAF-LOW. The recommended pair of [*unsw*, *nk*] can be used in Python. If using MATLAB only then [*unsw*, *rdeco*] is recommended as it provided coverages of *≥*80% on TELE and SAFER. However, this pair is a compromise in comparison to [*unsw*, *nk*], as it resulted in much less accurate RR-intervals on SAFER, and almost twice as many RR-intervals being unavailable for analysis on both datasets.

When requiring the highest possible accuracy regardless of the proportion of time for which RR-intervals are deemed suitable for analysis, we tentatively recommend [*unsw*, *jqrs*]. This combination uses two MATLAB QRS detectors. When using Python only, we tentatively recommend [*unsw*, *nab*]. We describe these recommendations as ‘tentative’, because these only achieved small improvements in accuracy in comparison to the pairs chosen when requiring accurate RR-intervals for a high proportion of the time. For instance, [*unsw*, *jqrs*] achieved an MAE in RR-intervals of 16.1ms and 13.1ms on TELE and SAFER respectively, compared to 19.8 and 16.3ms on TELE and SAFER for the previously recommended [*unsw*, *nk*]. This small improvement was achieved at the expense of making RR-intervals available for analysis for only 63 and 62% of the time on TELE and SAFER, compared to 89% for [*unsw*, *nk*]. Similarly, [*unsw*, *nab*] achieved MAEs of 21.5 and 14.7ms on TELE and SAFER, which represent was a debatable improvement compared to [*unsw*, *nk*]. Therefore, we expect the most useful QRS detector pairs will be those recommended for obtaining accurate RR-intervals for a high proportion of the time.

Future work should assess whether the best-performing QRS detector pairs continue to perform well across additional datasets and different recording scenarios.

### 4.3. Comparison with Literature

The importance of signal quality assessment techniques in the analysis of telehealth ECG signals has been well-documented in the literature. The use of signal quality assessment algorithms to exclude low-quality ECG signals from manual review during ECG-based AF screening has been proposed as a strategy to: (i) reduce cardiologist reviewing workload [38, 39]; (ii) improve the reliability of cardiologist diagnoses [40]; and (iii) improve the performance of automated AF detection algorithms [41]. Indeed, such algorithms are now commercially available and in use in large trials [42]. Whilst there has been much work on developing ECG signal quality assessment techniques [43], less work has focused on developing techniques specifically to assess whether RR-intervals can be accurately extracted from ECGs. Herein lies the main novelty of this study: we designed a novel algorithm to assess whether RR-intervals can be accurately extracted, and assessed its performance on real-world telehealth ECGs. This is valuable because the extraction of RR-intervals is a foundational step in several ECG analyses.

This work extends previous research into QRS detection and ECG signal quality assessment. We used the QRS detection algorithms which have previously been identified as the best-performing in telehealth settings in [12, 44]. We employed these in an adaptation of the previously proposed *bSQI* signal quality assessment approach [31]. The *bSQI* approach has previously been used to assess signal quality on windows of 1s duration [36] and 10s duration [28, 32]. In this work, we adapted the *bSQI* approach to assess whether individual RR-intervals had been accurately extracted. We believe our recommendation of the [*unsw*, *nk*] QRS detector pair is novel since to our knowledge previous work has not considered the *nk* QRS detector in *bSQI* algorithms [28]. We have extended our own previous work [22] by assessing algorithm performance on multiple datasets, investigating the impact of arrhythmia and noise on performance, and refining the duration of tolerance window used for signal quality assessment. This has led to a more informed recommendation of the [*unsw*, *nk*] QRS detector pair, and increased confidence about its generalisability.

One shortcoming of the proposed approach is that it produced less accurate RR-intervals on low quality ECGs. However, it should be noted that the low quality ECGs used in this assessment represent the lowest quality ECGs encountered in AF screening, extracted from the 2.8% of ECGs identified as poor quality in the SAFER Feasibility Study [38]. Therefore, this shortcoming will only be encountered for a small minority of real-world ECGs.

The proposed approach to assessing which RR-intervals are likely to be accurate relies on a tolerance window - the maximum permissible difference in the timings of QRS complex detections between the two QRS detectors. In this study we used a tolerance window of 100ms, which differs from the 150ms used in previous studies [22, 28]. The choice of tolerance window duration is significant for two reasons. First, a longer tolerance window tends to increase the proportion of time for which RR-intervals are assessed to be accurate, because there is greater tolerance of differences in QRS detections. Second, a longer tolerance window tends to result in a higher mean MAE in RR-intervals, because QRS complexes do not need to be detected as accurately for RR-intervals to be deemed suitable for analysis. The choice of 100ms was based on a preliminary analysis of performance when using different tolerance window durations, striking a balance between maintaining a high mean coverage and relatively low mean MAE.

### 4.4. Strengths and Limitations

The key strengths of this study are as follows. First, algorithm performance was assessed on real-world telehealth ECG signals recorded by patients at home. Second, we used open-source QRS detection algorithms, allowing others to implement the proposed approach in their own applications. Third, we have made the code used in this study publicly available. The analysis code is available at https://github.com/sharonho1215/ecg-signal-quality/, ensuring others can reproduce the study. A Python implementation of *unsw* is available at https://github.com/peterhcharlton/unsw_python/, which was shown to provide similar results to the original MATLAB version. This ensures that our recommended [*unsw*, *nk*] pair is in a single language (Python), and thus has greater utility. Fourth, the proposed algorithm has been implemented in the open-source Neurokit2 Python Package [25].

The key limitations of this study are as follows. First, performance was assessed on data collected from handheld devices at rest. It may be prudent to investigate performance in additional telehealth settings, such as with wrist-worn or chest-strap devices, rather than assuming that the tool will generalise to such settings. Second, the *unsw* QRS detector was developed using the TELE dataset [14]. For this reason, we have placed greater emphasis on results obtained using the SAFER dataset. Third, we did not compare the proposed tool with alternative ECG signal quality assessment techniques (such as [20, 39] - see [43] for a review), or combine it with other techniques to improve performance [32, 21, 16]. We also did not assess the utility of the tool for tasks other than RR-interval extraction, such as ECG signal quality assessment prior to extraction of other features. Fourth, we did not investigate whether the extracted RR-intervals are of sufficient accuracy for purposes such as heart rate variability analysis. Finally, the proposed tool is based on the assumption that when two QRS detectors agree on the timing of QRS complexes, then it is likely that the detected QRS complexes are accurate. Whilst the results indicate that this assumption is reasonable on high-quality ECGs, the results on low-quality ECGs indicate that it is not always valid.

### 4.5. Implications

This study reports a simple algorithm to accurately extract RR-intervals from telehealth ECGs. This algorithm could form part of a pipeline for automated analysis of single-lead ECGs recorded using devices such as handheld ECG recorders and smartwatches. Potential applications include detecting AF in daily life [5], and potentially identifying signs of other heart conditions such as supraventricular tachycardia [45]. In these contexts, algorithms could be used to: (i) identify ECGs which show signs of pathology, and safely exclude the remainder from clinical review [7]; (ii) make preliminary ECG interpretations for confirmation in clinical review [46]; and (iii) prioritise ECGs for clinical review according to their likelihood of showing pathology [47].

Future work should investigate whether the extracted RR-intervals are sufficiently accurate for particular use cases. In this study the extracted RR-intervals were found to be similarly accurate during AF (MAE of 6.7ms in AF compared to 7.7ms in non-AF), suggesting they may well be suitable for AF detection. It is not yet clear whether the extracted RR-intervals remain accurate at elevated heart rates such as those encountered in supraventricular tachycardia. Nor is it clear whether the extracted RR-intervals would be sufficiently accurate for any heart rate variability analyses. Therefore, further work is required to assess the clinical utility of the proposed algorithm for particular use cases, and to evaluate whether the increased accuracy provided by the algorithm improves performance on downstream tasks.

Future work may also investigate improving the proposed algorithm’s performance and robustness to noise. Performance could potentially be improved by incorporating information from more than two QRS detectors into the assessment of RR-interval accuracy through straightforward voting, or weighting information through confidence metrics. This is a similar approach to that presented by Liu *et al.*, who incorporated information from up to eight QRS detectors in an ECG quality assessment algorithm [28]. Performance could also be improved by combining information from QRS detectors with quality metrics derived from the underlying signal via machine learning [16]. The algorithm’s robustness to noise could potentially be increased by utilising advanced filtering techniques to eliminate noise from ECGs and their derivatives prior to QRS detection [48, 49, 50]; or utilising deep learning for improved QRS detection in the presence of noise [51].

The proposed algorithm may also have utility in real-time ECG signal quality assessment. Since the algorithm is relatively efficient (see [12] for QRS detector execution time assessments), it could potentially be used in near real-time to assess the proportion of an ECG from which RR-intervals can be accurately extracted. The user could then be prompted to finish recording if sufficient high quality data had been obtained, or to continue recording to obtain more data.

The characteristics of optimal QRS detector pairs varied according to use case. In some use cases such as AF screening, multiple ECGs are available per patient, presenting the possibility of discarding more data to prioritise even more accurate RR-intervals. Considering this use case: accurate RR-intervals were obtained for the highest proportion of time when using a pair of the most accurate QRS detectors (*e.g. unsw*, *nk*, *two-avg*, or *rdeco*, all of which performed reasonably well in our previous benchmarking study [12]). In some use cases only a single ECG may be available, making it important to extract accurate RR-intervals from a high proportion of the available data. For this use case, the most accurate RR-intervals were obtained, albeit for a smaller proportion of the time, when pairing a more accurate QRS detector with a less accurate QRS detector. This pairing is more consistent with the original design of the *bSQI* approach, in which a pair of QRS detectors was chosen where one was less sensitive to noise, and the other more sensitive to noise [32]. In contrast, in this study we found that obtaining accurate RR-intervals for a higher proportion of time requires two QRS detectors which are more robust to noise, diverging from the original *bSQI* approach. This is particularly important when analysing telehealth ECGs, which are more prone to noise. Consequently, this study adds to our understanding of how to select optimal QRS detectors for the *bSQI* approach.

## 5. Conclusion

In this study we developed an algorithm to accurately extract RR-intervals from single-lead, telehealth ECGs collected by home-dwelling patients using hand-held devices. The algorithm uses a primary QRS detector for RR-interval extraction, and a secondary QRS detector to assess which RR-intervals have been extracted accurately and therefore should be included in analysis. Based on results from two datasets, we recommend using the [*unsw*, *nk*] pair of QRS detectors. This produced mean MAEs in RR-intervals of 19.8 and 16.3ms on TELE and SAFER datasets respectively, and assessed 89% of RR-intervals to be accurate on both datasets. These MAEs were lower than when using either *unsw* or *nk* alone without quality assessment, demonstrating the benefit of using this algorithm for quality assessment. The accuracy of the RR-intervals derived using this algorithm was similar between AF and non-AF, but lower on low quality signals.

## Data Availability

The TELE ECG Database is publicly available at https://doi.org/10.7910/DVN/QTG0EP. Requests for access to the SAFER dataset should be directed to the SAFER study coordinator (SAFER @medschl.cam.ac.uk) and will be considered by the investigators, in accordance with participant consent.

## Acknowledgment

This study is funded by the British Heart Foundation (FS/20/20/34626 awarded to PHC), the National Institute for Health and Care Research (NIHR) Programme Grants for Applied Research Programme (RP-PG0217-20007 awarded to JM), and the NIHR School for Primary Care Research (SPCR-2014-10043, project 410 awarded to JM), and by a W.D. Armstong Trust Fund studentship awarded to ZD. The views expressed are those of the author(s) and not necessarily those of the NIHR or the Department of Health and Social Care. The funders had no role in study design, data collection and analysis, decision to publish, or preparation of the manuscript.

## Appendix A. Selecting the tolerance window size

A tolerance window size of *±*100ms was chosen based on the preliminary analysis shown in Figure A.7(a). On the SAFER dataset it was found to provide a relatively high mean coverage, whilst also maintaining a relatively low mean MAE (see Figure A.7(b)). This selection was subjective, and any value between approximately 60 and 140 ms would have achieved similar results.

**Figure A.7:**
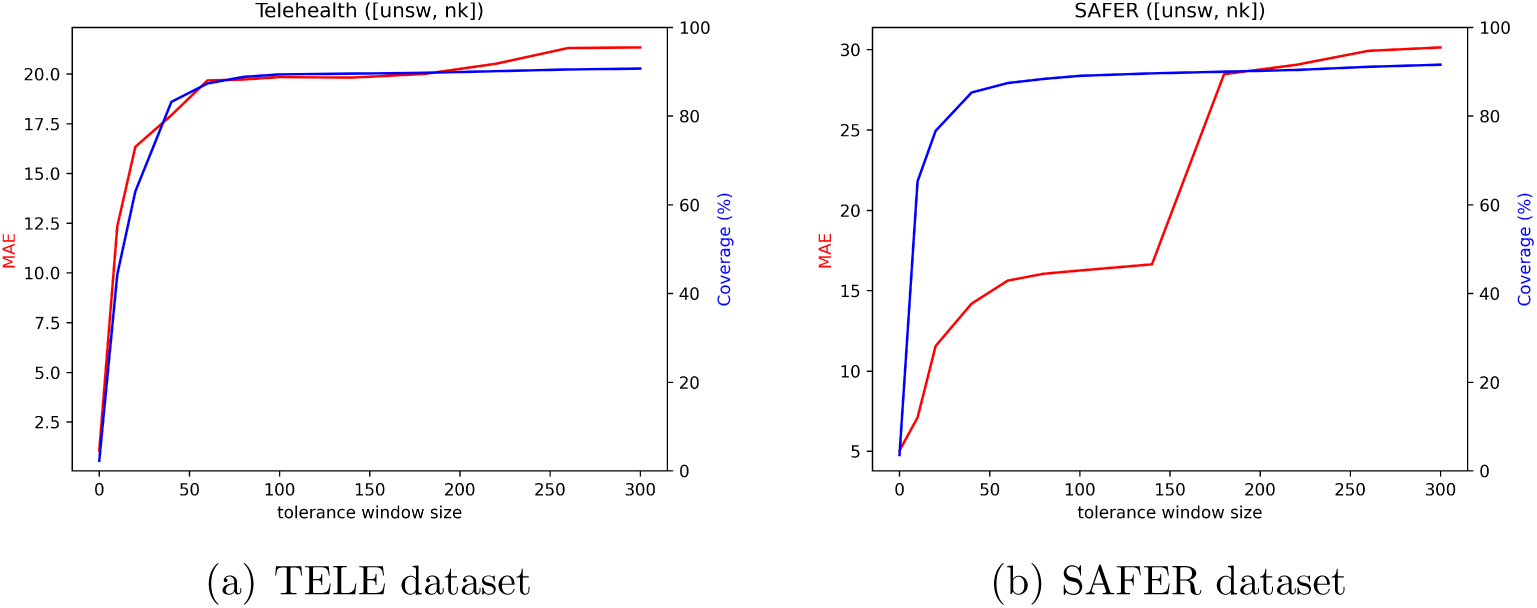
The effect of window size on performance. The plots show how mean MAE and mean coverage varied with different tolerance window sizes for the [*unsw*, *nk*] QRS detector pair.

## Appendix B. Results obtained using the original MATLAB implementation of *unsw*

The results reported in the main text were obtained using a new Python implementation of *unsw*. Here we report corresponding results obtained using the original MATLAB implementation. Results were similar when using MATLAB and Python implementations.

The overall performance when using the MATLAB implementation of *unsw* is shown in Figure B.8, which corresponds to the results obtained using the Python implementation of *unsw* in Figure 3. The differences between *unsw* implementations were minimal: differences in MAEs of *≤* 3ms on TELE and *≤* 6ms on SAFER (for all combinations except the outlier of [*unsw*, *gqrs*]); and differences in mean coverage of *≤* 1%.

Similarly, the performance on each of the SAFER subgroups when using the MATLAB implementation of *unsw* is shown in Figure B.9, which corresponds to the results obtained using the Python implementation of *unsw* in 5. On high-quality ECGs, the differences between *unsw* implementations were again small: differences in MAEs of *<*5ms on high-quality ECGs (SAFER AF-HIGH and SAFER nonAF-HIGH), and *<*6ms on low-quality ECGs (SAFER nonAF-LOW) (for all combinations except the outlier of [*unsw*, *gqrs*]); and differences in mean coverage of *≤* 1%.

**Figure B.8:**
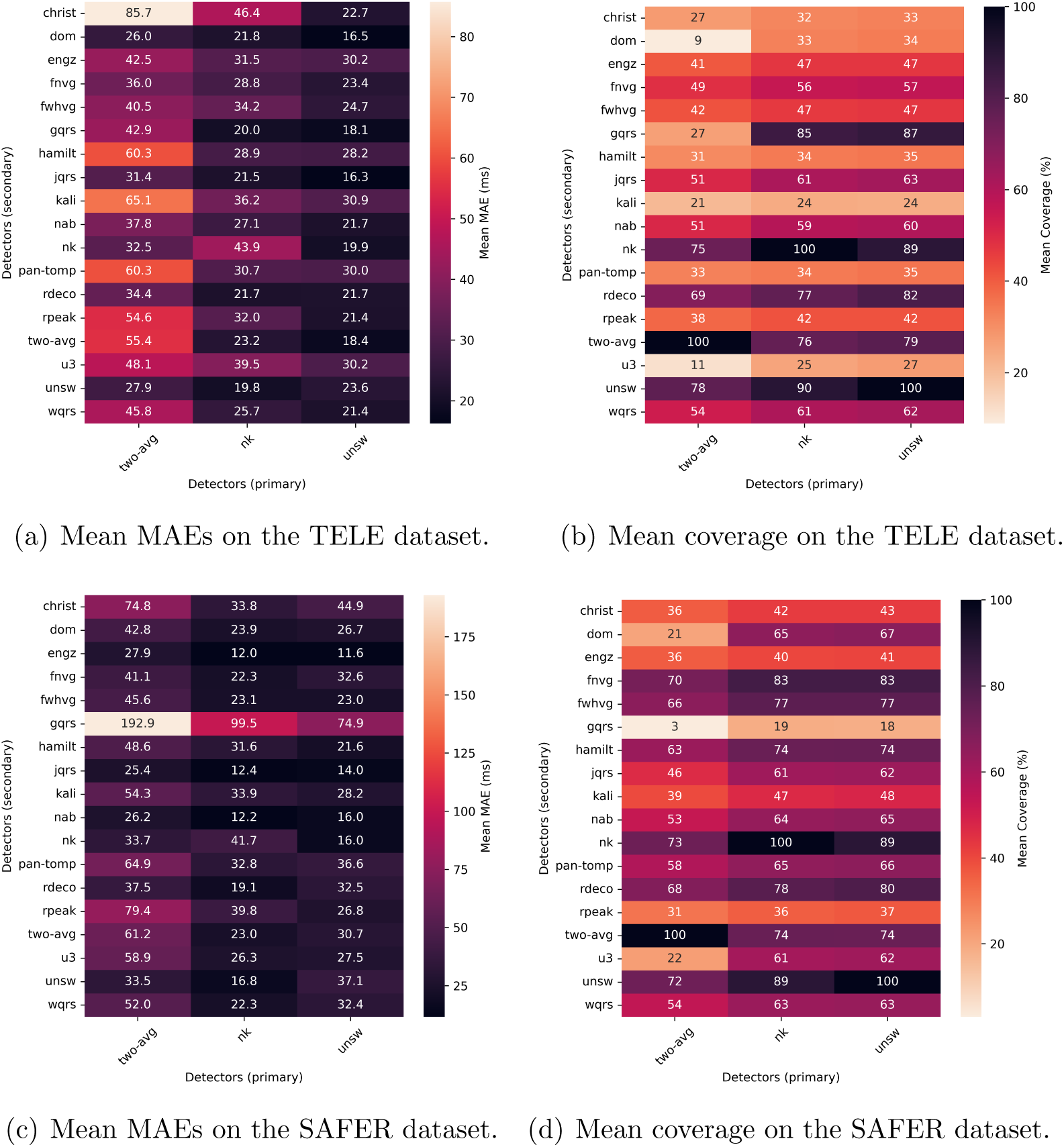
Overall performance when using the MATLAB implementation of *unsw*. Heatmaps showing the mean MAE ((a) and (c)) and mean coverage ((b) and (d)) for each pair of QRS detectors on the TELE ((a) and (b)) and SAFER ((c) and (d)) datasets. Here, the ‘coverage’ is the proportion of time for which RR-intervals were assessed to be accurate.

**Figure B.9:**
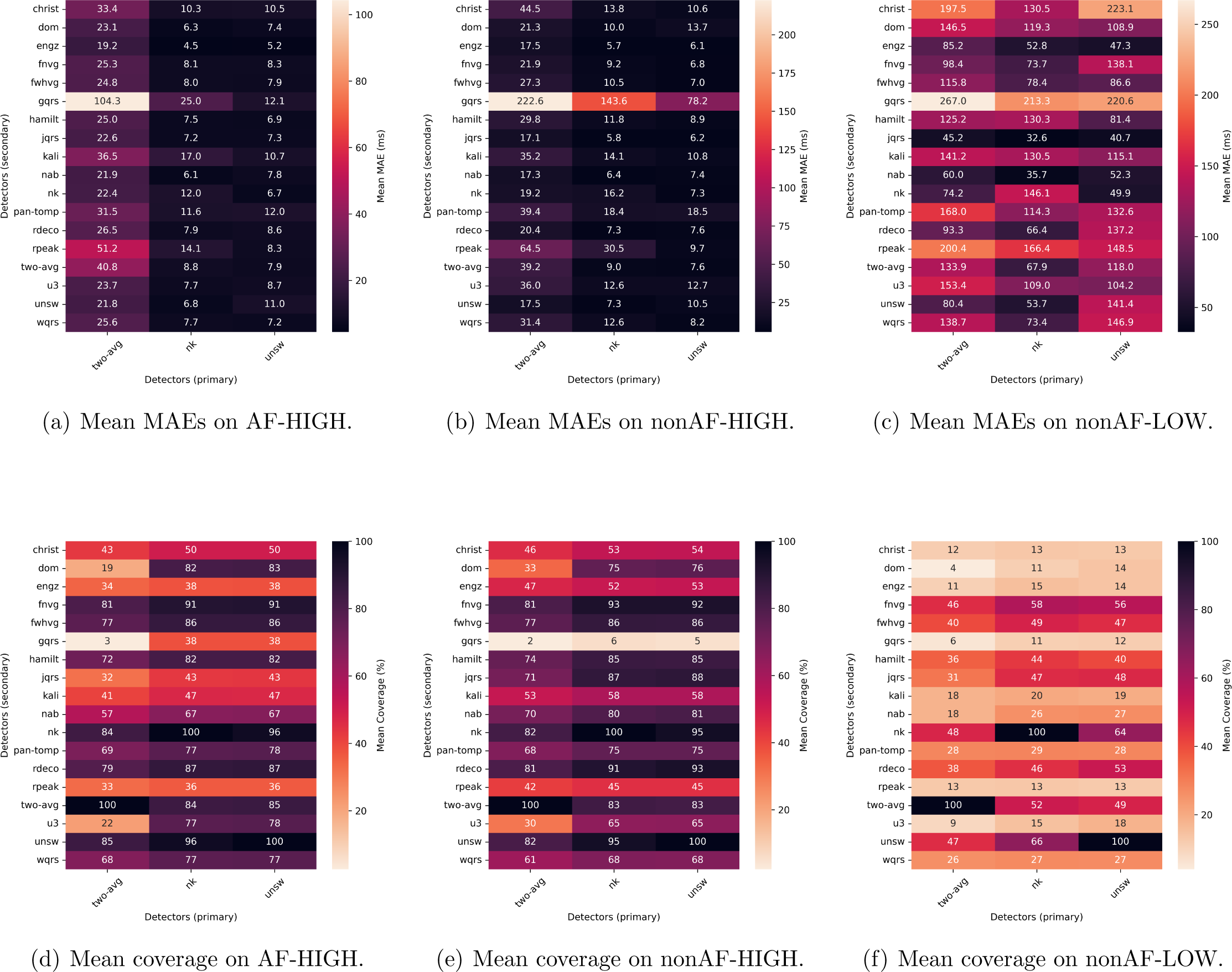
Comparing performance across SAFER subgroups when using the MATLAB implementation of *unsw*. Heatmaps showing the mean MAE ((a), (b) and (c) and mean coverage ((d), (e) and (f)) for each pair of QRS detectors on the SAFER AF-HIGH ((a) and (d)), SAFER nonAF-HIGH ((b) and (e)), and SAFER nonAF-LOW ((c) and (f)) subgroups.

